# Effectiveness of mHealth Interventions for Blood Pressure Control among Vulnerable Populations: A Systematic Review

**DOI:** 10.1101/2023.04.07.23288278

**Authors:** Kristen M.J. Azar, Yael Zoken, Rhonda M. Cooper-DeHoff, Valy Fontil, F. Modrow Madelaine, Catherine Nasrallah, Mark J. Pletcher

**Author notes:** Corresponding author: Kristen M.J. Azar, RN, MSN/MPH.

## Abstract

Social determinants of health and sociodemographic differences place some individuals at higher risk for hypertension leading to persisting disparities. While mobile health (mHealth) offers a promising approach to facilitate blood pressure (BP) management, it remains unclear which interventions are most effective for addressing disparities in hypertension control. We reviewed the current literature to examine whether mHealth interventions for BP control are effective in improving BP control in populations vulnerable to disparities in hypertension. We conducted a systematic review using multiple databases from January 1, 2009 through December 31, 2020. For inclusion, participants must have elevated blood pressure and belong to a sociodemographic group with known disparities in hypertension. We also tracked specific characteristics of each intervention. Out of the eight articles that met our eligibility criteria for inclusion, five were published in 2018 to 2020. Only four demonstrated a significant reduction in BP and all those interventions incorporated care teams. Despite some evidence of the effectiveness of mHealth interventions for improving BP control among vulnerable groups, more effective interventions are needed, and the quality of studies is overall low. Further research is needed to find the most effective ways to engage diverse communities with mHealth solutions to improve BP control.

## Introduction

Uncontrolled blood pressure (BP) causes over 450,000 deaths per year in the United States (US), making it a leading preventable cause of death.^1,2^ While national data indicate that BP control rates in the U.S. have improved over time, from approximately 30% in the early 1990’s to 50% in more recent years,^3,4^ a recent study published in the Journal of the American Medical Association revealed that HTN control has declined to 43.7% in 2017-2019, and further among minoritized groups.^3^ Further, race/ethnic and socioeconomic disparities exist, with African American (AA) individuals and those with low-income status being at higher risk for uncontrolled HTN.^5^ In communities with low socioeconomic and resource, the adverse cardiovascular outcomes of HTN and challenges of self-management are more severe due to social determinants such as low health literacy, lack of access to quality health care, and lack of trust in health care professionals and institutions.^6^

A growing body of evidence supports the use of mobile health (mHealth) interventions for health promotion, behavior change, diagnosis, and self-management of risk factors for chronic HTN. MHealth can be defined as “medical and public health practice supported by mobile devices such as mobile phones, patient monitoring devices, personal digital assistants, and other wireless devices”.^7,8^ It is estimated that more than 90% of U.S. adults own a mobile phone, including 83% AA and 76% of individuals with a low with low-income status, and approximately 85% have a smartphone.^9^ The use of mHealth interventions is increasing substantially in the general population given the near ubiquity of smartphone and mobile health technologies in all segments of society.^10^ The integration of mHealth technology (i.e. home BP monitoring devices and smartphone apps) with electronic health record (EHR) data has the potential to provide a major mechanism for interventions for BP management. It is becoming increasingly feasible to help patients self-manage their HTN remotely, allow patients to engage and benefit from the comfort of their home environments and communities. Self-management tools are designed to facilitate autonomy in self-management, augment patient/clinician collaboration, and to enhance patient empowerment.^11^

While mHealth has been shown to be effective in reducing CVD risk for affluent and well-resourced patient populations^12,13^, it is unclear how effective these interventions are among underserved and socioeconomically vulnerable populations, given existing health and healthcare disparities. Several studies provide evidence of the positive impact of mHealth on patient adherence with HTN medication regimens.^14^ and there is limited evidence to support the potential for mHealth to improve HTN outcomes among vulnerable populations, albeit the evidence is limited in part to a dearth of interventions specifically targeted to these populations. The objective of this study is to describe the characteristics of mHealth interventions and to examine whether these interventions are effective for reducing systolic and/ or diastolic blood pressure for socioeconomically vulnerable populations (e.g. socioeconomic or race/ethnic groups shown to have higher prevalence of hypertension).

## Materials and Methods

### Review Design and Study Selection

We conducted a systematic review to assess effectiveness of mHealth interventions for BP management among individuals with elevated BP in socioeconomically vulnerable populations. This review was registered in advance on PROSPERO under study number CRD42020169115.^15^ We followed the Preferred Reporting Items for Systematic Reviews and Meta-Analyses (PRISMA) guidelines for conducting and reporting items for systematic reviews.^16^

We used the PICOS terminology to frame this work. Participants are those with suboptimal BP who belong to a socioeconomic group with known disparities in hypertension. Interventions of interest are experimental or quasi-experimental mHealth intervention studies to reduce BP. The main comparators are usual care or other non-mHealth interventions for BP management. The primary outcome of interest is reduction in systolic and/or diastolic BP.

### Search Strategy

We searched the following databases for primary studies: MEDLINE® via PubMed, PsychInfo, CINAHL® and EMBASE from January 1, 2009, through December 31, 2020. We limited the search to studies occurring after 2009 given the significant advances in mHealth technology over the past decade. We developed a search strategy for MEDLINE, accessed via PubMed®, based on medical subject headings (MeSH®) terms and text words of key articles that we identified a priori (Appendix 1). Our basic search concepts were “mHealth AND hypertension”. We first identified potentially eligible studies based on title and abstract. One reviewer (KMJA) reviewed the titles and abstracts based on pre-specified inclusion/exclusion criteria. Two reviewers (KMJA and YL) independently reviewed the papers identified for full text review using a standardized data extraction form electronically entered into a web-based survey tool (Appendix 2). The full text of the final studies were then reviewed by a third researcher (MP). All discrepancies were resolved by consensus.

### Study Selection

#### Study Characteristics

We included interventions that assessed BP self-management using mHealth in the U.S., Canada, Western Europe or Australia for hypertension management in our target population as defined below. Studies were included if they were of experimental or quasi-experimental design, assessed the primary outcome of reduction in systolic and/or diastolic BP at least 3 months post-baseline and were published in English. Studies were excluded if the primary outcome was specifically related to ocular hypertension or pulmonary hypertension. Studies with a primary outcome of medication adherence were excluded. Studies that were non-experimental in design were excluded, as were studies published only as abstracts.

#### Target population

Adults (age 18+) with evidence of elevated BP at baseline (i.e. documented evidence of systolic BP ≥ 130 mmHg and/or diastolic BP ≥80 mmHg) and meeting criteria for socioeconomic and racial/ethnic vulnerability relative to bearing disproportionate risk for uncontrolled HTN.^17^ Studies were included if study participants had < 50% non-Hispanic White, with > 50% high school education or less, with > 50% Medicaid or > 50% annual household income ≤ $50K, and/or a primarily and explicitly defined rural population. Studies were excluded if the patient population included individuals with a life-threatening co-morbid illness (i.e. cancer diagnosis, end stage renal or liver disease) or pregnant women.

#### Intervention Inclusion/Exclusion

We include technology-enabled or enhanced interventions that specifically utilize mHealth. The Global Observatory for eHealth of the World Health Organization defines mHealth as “medical and public health practice supported by mobile devices, such as mobile phones, patient monitoring devices, personal digital assistants, and other wireless devices”,^18^ or the use of these technologies for health services and information.^19^ Given this definition, eligible interventions must include wearables and/or devices connected to an app, mobile site and/or mobile internet via wireless connection (e.g. Wi-Fi, blue tooth etc.). The intervention must be intended to reduce systolic and/or diastolic BP as the primary outcome of interest. Additionally, the intervention must be remote (i.e. used outside of the clinic setting) and patient-facing (i.e. patient interfaces and uses the technology outside of the clinic setting), but may include a clinic-based component. We also excluded studies with interventions requiring a surgical or invasive component (i.e. implanted or internal BP monitor).

### Data Extraction and Quality (Risk of Bias) Assessment

Data were systematically extracted and summarized. We reported study characteristics and intervention characteristics, using categories to describe components of included studies to facilitate cross-study comparisons. Primary outcomes were reported for all studies. Secondary outcomes (i.e. medication adherence) were reported for studies that included these outcomes using the metric utilized by the original study.

All three researchers (KMJA, YL and MP) independently assessed the risk of bias of the eight included studies using the Cochrane Risk of Bias tool. The specific domains include random sequence generation, allocation concealment, blinding, incomplete outcome data, selective reporting, and other source of bias. Any disagreement will be resolved by discussion between the three reviewers.

## Results

### Identification of Studies and Study Selection

We identified 2,574 articles through the literature search. After excluding duplicates, 2,023 articles were screened for inclusion based on title and abstract. Of these, 255 abstracts were review by one research (KJA) to identify manuscripts requiring a full text review. Two researchers (KJA & YFZ) independently assessed 74 full-text papers for eligibility and both authors agreed to exclude 66 articles based on the established protocol and criteria. The remaining 8 articles were identified as meeting the eligibility criteria for inclusion and were then reviewed by a third researcher (MP) for confirmation. Reasons for exclusion of the 66 full-text articles are indicated by the PRISMA diagram (Figure 1).

**Figure 1:**
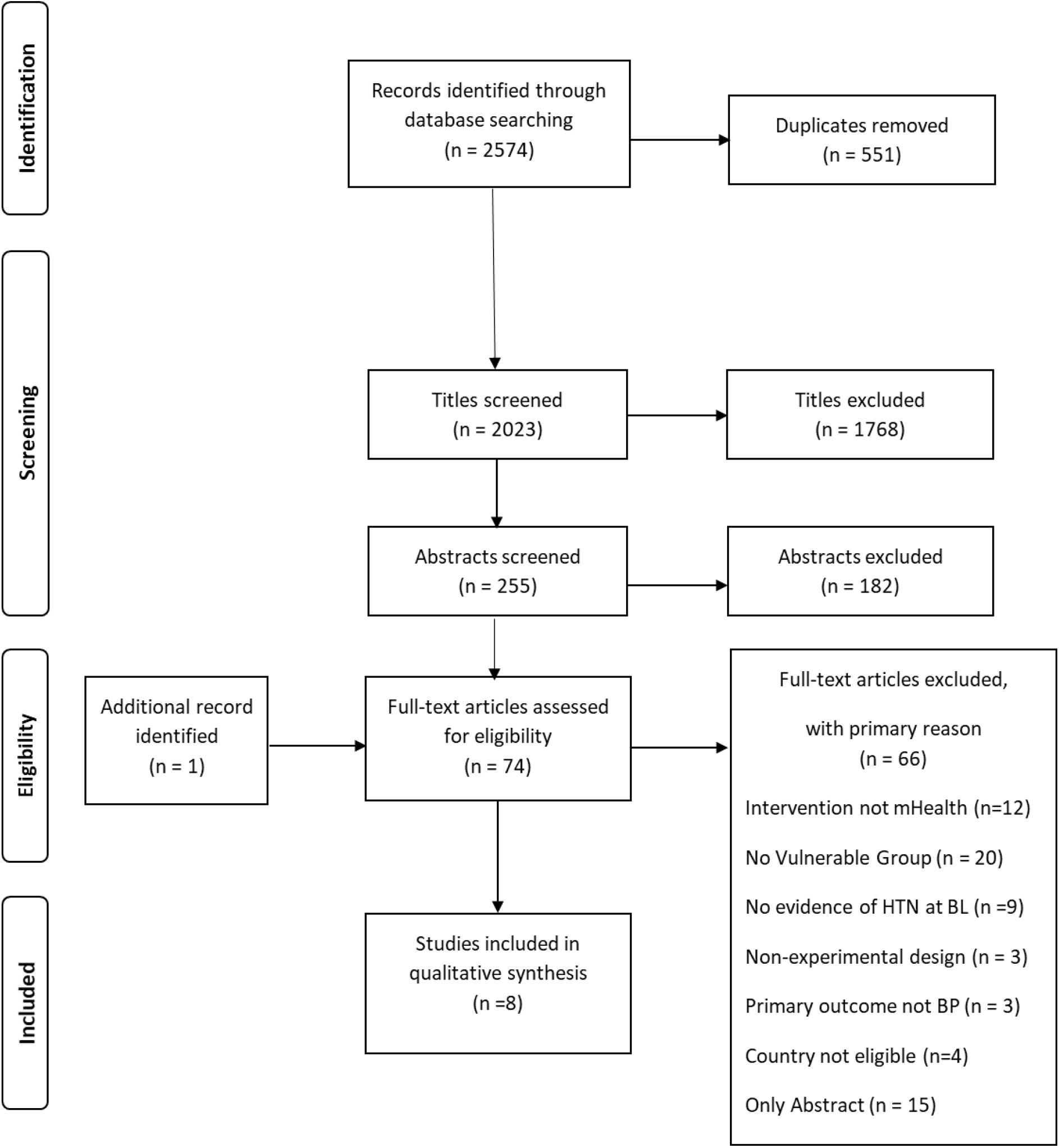
PRISMA Diagram.

### Study Characteristics, Design and Setting

Characteristics of the eight studies are presented in Table 1. Of the eight studies identified for inclusion, six were published since 2017, six were conducted in the United States, one in the United Kingdom^20^ and one in Canada.^21^ All eight studies were randomized control trials with follow-up ranging from 3 to 12 months. Half of the studies compared the mHealth intervention to usual care^20,22-24^, three used an enhanced usual/standard care group and one involved a passive intervention as the comparison group.^21^

**Table 1:**
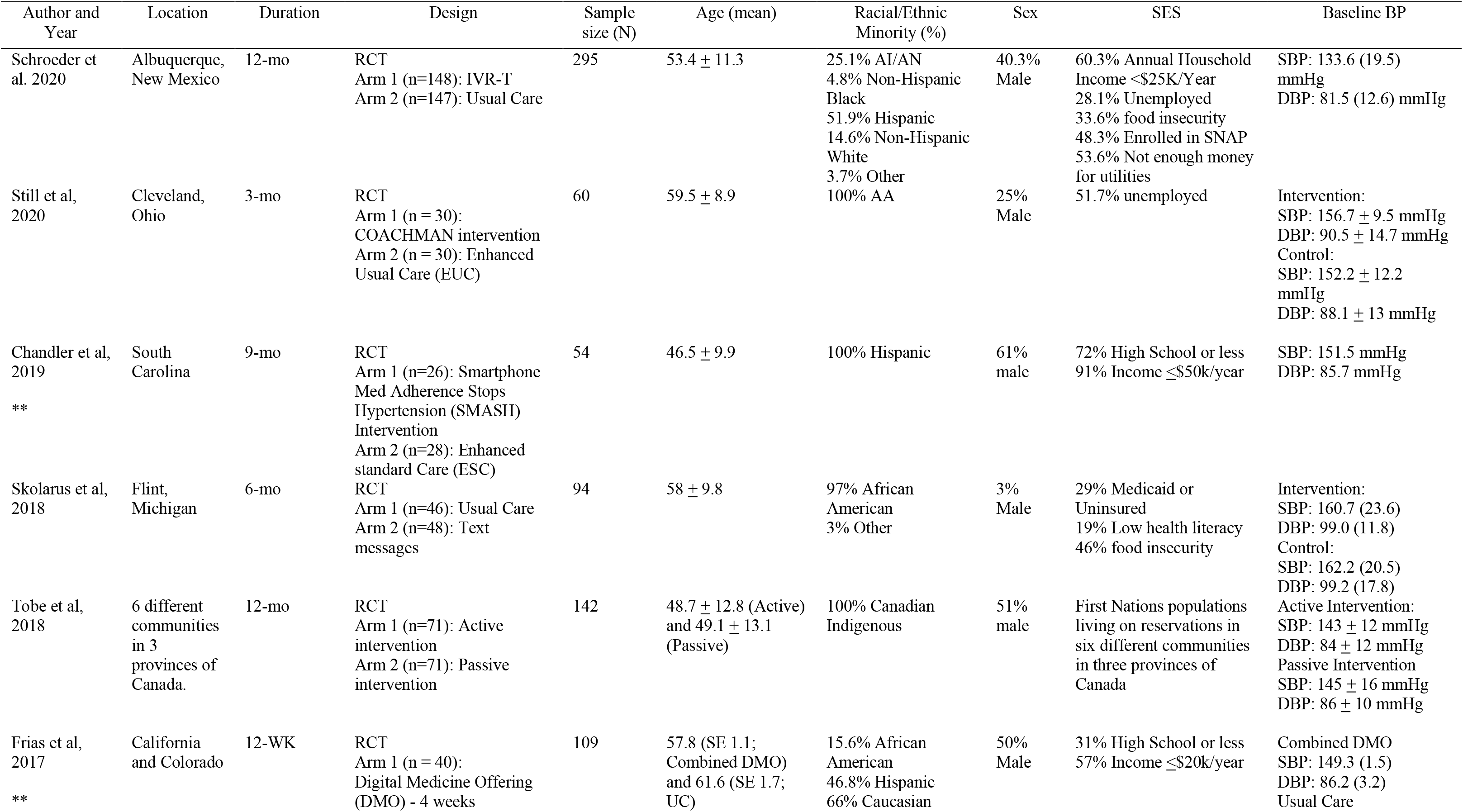
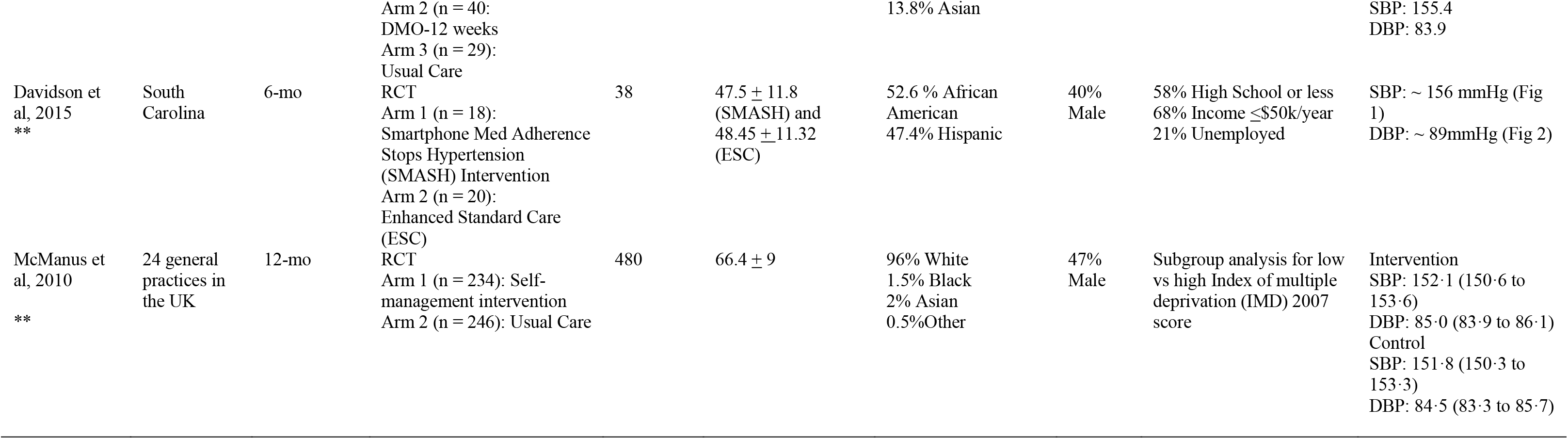
Study Design, Quality, and Participant Characteristics

#### Participant characteristics

The number of participants in a given study ranged between 38 and 480 individuals. For three studies, at least 50% of participants were male. Among the studies, three^23,25,26^ had more than 50% Non-Hispanic Black participants. One study included only Hispanic participants^27^ and another included only Canadian Indigenous groups.^21^. In four studies^22,24,25,27^ at least half of the participants had an annual income of ≤ $50K. One study^20^ conducted an a priori secondary analysis for low vs high scores for the Index of Multiple Deprivation. All but one study^24^ included participants with a baseline BP in the Hypertension Stage 2 range (SBP ≥ 140 mmHg and/or diastolic DBP ≥90 mmHg).^17^

### Intervention Characteristics

We examined mHealth intervention characteristics across the eight included studies (Table 2). The eight studies included seven unique interventions, given that two studies^25,27^ tested the same intervention (i.e. Smartphone Medication Adherence Stops Hypertension [SMASH]). Five interventions were developed with direct involvement from members of a targeted vulnerable community.^21,23-27^ More than half (n = 4; 57%) of the interventions involved the use of a smartphone and/or smartphone application (app).^21,22,25-27^ Five interventions (71%) required remote self-monitoring with an automated sphygmomanometer (arm cuff).^20,21,23,25-27^ While one study^24^ did not require use of an automated sphygmomanometer, the option was offered and 98% of participants did use one. One study^22^ utilized an ingestible sensor placed in a pill, with a wearable sensor patch intended to promote medication adherence. Three interventions offered educational content.^21,24,26^ Three interventions used SMS and text messaging^21,23,25,27^ to deliver tailored motivational messages, educational content and /or adherence reminders via mobile phone. Three offered visualizations for self-monitored BP metrics and longitudinal trends, adherence and tracking of other health behaviors.^20,22,25,27^ Most interventions (n = 5; 71%) ^20-22,25-27^ involved a clinical care team, which received transmission of data and/or provided supplemental counseling.^26^

**Table 2:**
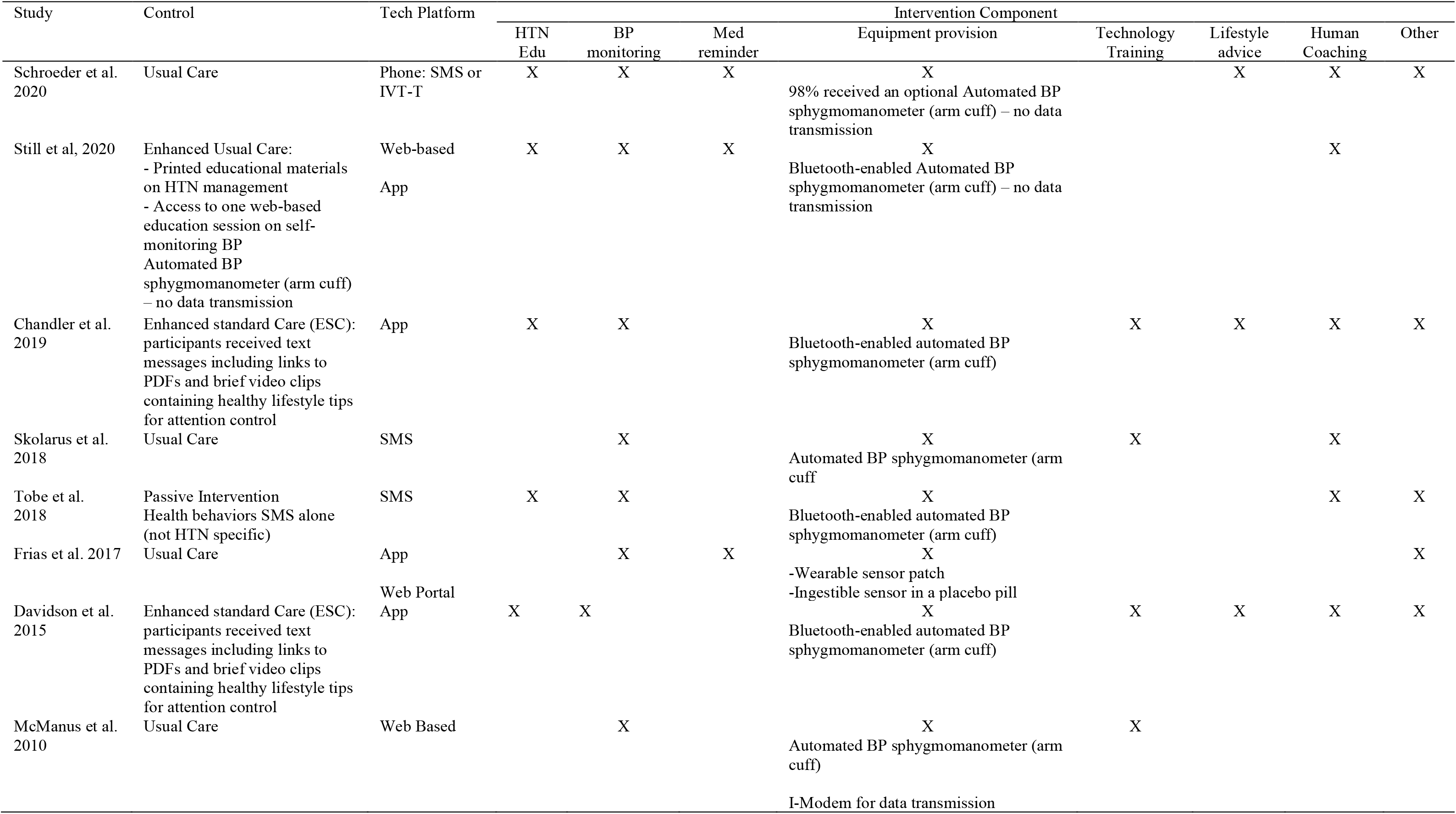
. Intervention Features and Components

### Intervention Effectiveness / Results of Individual Studies

Four studies^20,22,25,27^ (including three different interventions) demonstrated a significant reduction in mean SBP and/or DBP for treatment vs comparison group participants (Table 1). Davidson et al (2015) and Chandler et al (2019) tested the SMASH intervention whereby adult participant were randomly assigned to either the SMASH group or the Enhanced standard care (ESC) group. The intervention was tested in two randomized controlled trials, one largely composed of low socioeconomic status (SES) Hispanic adults (n = 54)^27^ and the other with low SES Hispanic and AA adults (n = 38, 53% AA and 47%Hispanic,).^25^ The intervention led to a reduction of the SBP in the SMASH group. The SMASH program involves a smartphone app to improve BP medication adherence as well as Bluetooth-enabled automated BP device. Participants were asked to measure their BP using the device and app every three days. It also facilitates timely patient-provider communication and included tailored reinforcement/motivational messages via SMS. The development of the intervention was guided by Self Determination Theory^28,29^ and aimed to increase autonomous motivation by linking participants’ behavioral changes to their values, beliefs and goals. Messages are based upon participant responses to a branch logic questionnaire that identified individual values, beliefs and short/long-term goals. The 600+ messages were developed using formative research methods among the target population (i.e. Hispanic adults with hypertension). The intervention also utilized the global systems for mobile electronic tray Maya (MedMinder, Inc., Newton, MA, USA), which provided a series of reminder signals to take medications. The data was shared with the care team to inform clinical care.

Frias et al (2017) conducted a three-arm randomized controlled trial to test the effectiveness of a digital medicine offering (DMO) among low SES adults with both type 2 diabetes and hypertension (n = 109) and found at 4 weeks compared to usual care. The intervention consisted of the use of an ingestible sensor co-capsulated with antihypertensive medication and transmitting data to an external sensor patch for medication adherence tracking.

Participants used DMO (includes digital medicines, the wearable sensor patch, and the mobile device app) for 4 or 12 weeks (treatment groups 1 and 2 respectively) or received usual care (treatment group 3). DMO reports were provided to clinicians to guide decision making for medication titration. While the trial was 12-weeks, the primary outcome was mean change in SBP at 4 weeks and was found to be significantly reduced in both treatment arms compared to usual care (Table 1) and maintained at 12 weeks. Also of note, providers taking care of a patient who was randomized to DMO intervention made approximately 3 times more medical decisions per participant compared to usual care.

McManus et al (2010) RCT (n = 480) tested the telemonitoring and self-management in the Control of Hypertension (TASMINH2) intervention and found change in mean SBP between baseline 6 and 12 months (SBP decreased by 17.6 mm Hg in the self-management group and by 12.2 mm Hg in the control group). The TASMINH2 intervention combined self-management of BP, self-titration of antihypertensive medications combined with telemonitoring of home BP measurements by the care team. Monthly summaries of the self-management data were sent to the clinical team for review and use in informing clinical care. While the intervention was web-based, it involved the passive transmission of data from the automated sphygmomanometer via i-Modem, inserted into a home electrical outlet. The analysis included a priori-defined subgroup analysis among those with low vs high Index of multiple deprivation (IMD) 2007 score. The overall reduction in SBP was greater for participants with low IMD compared with participants with high IMD 2007 at both 6 and 12 months (–0.4 mm Hg; 95% CI –5.9 to 5.2, p=0.05and 1.6 mm Hg; 95% CI –4.4 to 7.6, p=0.08 respectively). The remaining four studies^21,23,24,26^ did not demonstrate a significant reduction in SBP and/or DBP for the treatment vs comparison group participants. Schroeder et al (2020), tested the impact of an interactive voice response and text message (IVR-T) based intervention, compared with usual care, to deliver automated appointment and medication refill reminders as well as weekly culturally tailored (i.e. AI/NA) motivational messages to encourage self-management. The authors speculate that the null findings may in part be due to study design and a large proportion of participants with hypertension being in control at baseline. Both the Skolarus et al (2018) and Still et al (2020) interventions were developed in collaboration with direct involvement from members of the African American community, using the principles of community-based participatory research. One intervention^23^ consisted of prompted remote BP self-monitoring, tailored text-messages that are responsive to monitored data as well as both tailored and generic automated text-messages to promote healthy behaviors. The authors speculate that small sample size and low-intensity of the intervention may have contributed to null results. The other intervention^30^ combined self-directed web-based hypertension education, remote self-monitoring and app-supported medication management along with up to 4 patient-initiated, informal nurse counseling. They speculate that the provision of home BP self-monitoring devices to both groups could have resulted in greater awareness and behavior change for all study participants. Finally, Tobe et al (2019) tested a SMS text-based intervention among members of the Canadian Indigenous Groups community with hypertension. The intervention consisted of “active” messages to promote BP self-management with “passive” messages to promote generic healthy lifestyle behaviors, compared to passive alone.

### Strength of Body of Evidence, Risk of Bias and Quality of Studies

A summary of study quality, assessed via the Cochrane Risk of Bias Tool, is presented in Table 4. No studies that met the inclusion criteria were excluded from the review on the basis of quality. One^21^ of the 8 studies was rated high risk for both incomplete outcome data and selective outcome reporting. The risk of bias for these domains for the other studies was either unknown^23,26^ or low^20,22,24,27,31^. There was high or unknown risk of bias due to blinding participants & personnel for all of the studies.

## Discussion

In this systematic review we focused on mHealth interventions for BP reduction among individuals with hypertension and belonging to socioeconomically vulnerable and high-risk populations. We identified eight trials that met our eligibility criteria for inclusion. Four of these studies, which incorporated care team impact, demonstrated a statistically significant reduction in systolic BP.

While there have been several related systematic reviews conducted in recent years^32-39^, our review includes relatively recent literature, where five of the eight studies included were published in 2018 to 2020. One other recent review by Khoong et al.^40^ examined the impact of mobile health strategies for BP self-management on BP outcomes but the search was limited to studies conducted prior to July 2019 and, thus, does not include several of those we have identified.^21,22,24,26,27^ Among the main findings was a need for increased diversity in clinical research studies as well as the use of implementation science frameworks to facilitate comparison between different multi-modal studies. Given the rapidly evolving technology, frequent updates to reviews are justified to keep pace with these advancements. Our review intentionally uses a broader definition of mHealth interventions for BP, where prior reviews have limited focus only on smartphone apps^32,36^ and/or SMS interventions^34^ Despite persistent disparities, there remains limited evidence to identify effective mHealth strategies that will benefit groups that bear a disproportionate burden of hypertension risk and its related sequalae.^41^

Overall, prior systematic reviews found mixed evidence for the effectiveness of mHealth interventions for hypertension management. A recent systematic review and meta-analysis by Li et al. (2020) aimed to measure the effectiveness of mHealth in improving the self-management of hypertension for adults and found a greater reduction in both SBP and DBP in the mHealth intervention groups compared with control groups, −3.78 mm Hg (P<.001; 95% CI −4.67 to −2.89) and −1.57 mm Hg (P<.001; 95% CI −2.28 to −0.86), respectively.

They found that high intensity of medication reminders, user-driven designs with customized 2-way communication between patients and physicians and multifaceted functions (i.e. SMS text messaging, linking BP monitoring devices to a Web-based system, apps etc.) were associated with effectiveness. In a meta-analysis of randomized controlled trials testing interactive mHealth interventions for BP management in adults (total n = 4271 participants), Lu et al (2019) found that the interventions were associated with significant changes in systolic BP and diastolic BP of −3.85 mmHg and −2.19 mm Hg respectively. While they found evidence of effectiveness, there was no reporting of sociodemographic characteristics of participants and only one study included^22^ was also included in the present review.

Consistent with prior reviews, a common element among the effective interventions in our review seemed to be intentional involvement and communication with the care team, aimed at informing clinical decision making and medication titration. The four interventions with null results did not include a formal mechanism for information sharing with the care team and were, in general, lower intensity (i.e. no care team involvement; passive engagement) in comparison to the SMASH, DMO and TASMINH2 interventions. Some early studies in home-based telemonitoring of BP, where remote self-monitored data was shared with the care team, resulted in under treatment and worse outcomes.^42-44^ More recent evidence suggests benefit when clinical guidelines are adjusted to recommend lower targets for home readings.^45,46^ The SMASH intervention^25,27^ involved the generation of a biweekly report from self-monitored BP data that was used to inform clinical care and incorporated commonly available, nearly ubiquitous technologies. It is especially promising given that the results were replicated in a second study, though both studies were small (n < 100) and potential for scalability is unclear. The Frias et al (2017) intervention used an ingestible sensor co-capsulated with antihypertensive medication, which is not a commonly available or widely used technology. The authors did not report cost data and it is unclear how feasible it would be to scale up the intervention. Additionally, the authors reported some incidence of gastrointestinal related adverse events, raising questions of safety and acceptability in a broader context. It is important to note that the TASMINH2 study was conducted only in the United Kingdom and was found effective among a group identified as low SES but in the context of socialized medicine. Given this, there may be differences when deploying among socioeconomically disadvantaged groups in the U.S.

While health information technology has been recognized as a potentially powerful tool in addressing racial/ethnic disparities among Black communities, it is recognized that high quality research to improve the scalability and sustainability of culturally appropriate interventions is needed.^47^ Further, deficiencies in existing interventions that do not promote equitable outcomes must be identified and mitigated.^48^ While community-based approaches, such as barbershop and faith-based programs, have shown promise^47,49^, both studies in our review^23,30^ which incorporated these types of approaches were not effective for reducing BP. This could in part be due to limitations in study design (i.e. insufficient power) and also due to the low intensity of the mHealth interventions themselves.

Several other studies exploring mHealth interventions specifically among socioeconomically vulnerable adults were identified in our review or prior systematic reviews but did not meet our inclusion criteria and are worth noting. Buis et al (2017)^14^ tested daily, automated text reminders sent at individually tailored times, among Black adults with uncontrolled hypertension and did not find significant differences in BP outcomes. This study was excluded due to limited follow-up. Lewinski et al (2019)^50^ recruited patients of low socioeconomic status from a federally qualified health center with both diabetes and uncontrolled hypertension. The intervention consisted of app and text messages and did not result in improved hypertension control and did not include a comparison group. Neither of these studies involved the care team or incorporated medication management. Taber et al (2018)^51^ aimed to assess the efficacy of a pharmacist-led, technology-aided, educational intervention and found modest systolic BP reduction among Black participants (−0.86 mmHg per month, p=0.026), which was not evident in non-Blacks (−0.13 mmHg per month, p=0.865). This study was excluded due to a lack of comparison group. The findings in these studies are consistent with the potential importance of care team involvement for interventions aimed at reducing disparities in hypertension.

## Conclusions

These findings compel the crucial and critical need for innovation and for the development of more effective interventions specifically to improve outcomes among those at the highest risk and in the most need. These findings also compel the urgent need for those developing mHealth interventions for hypertension control to engage high-risk and disproportionately burdened populations. Additionally, the evidence suggests that mHealth innovations should include more clinical decision support as opposed to self-management alone.

## Data Availability

The data used to support the findings of this study are available from the corresponding author upon request.

## Funding Details

The research reported in this publication was supported by Patient-Centered Outcomes Research Institute (PCORI) [PaCR-2017C2-8153]. The content is solely the responsibility of the authors and does not necessarily represent the official views of BPTrack/PCORI

## Competing Interests

Mark J. Pletcher, MD, MPH has received research grants from the Patient-Centered Outcomes Research Institute (PCORI). All the other authors declare that there is no conflict of interest regarding the publication of this paper.

## Acknowledgments

We would like to thank Evans Whitaker, MD, MLIS, and academic librarian for UCSF Health Science Library.

## Supplementary Materials

### Supplemental Material #1

| Section/topic | # | Checklist item | Reported on page # |
| --- | --- | --- | --- |
| TITLE |  |  |  |
| Title | 1 | Identify the report as a systematic review, meta-analysis, or both. | 1 |
| ABSTRACT |  |  |  |
| Structured summary | 2 | Provide a structured summary including, as applicable: background; objectives; data sources; study eligibility criteria, participants, and interventions; study appraisal and synthesis methods; results; limitations; conclusions and implications of key findings; systematic review registration number. | 3 |
| INTRODUCTION |  |  |  |
| Rationale | 3 | Describe the rationale for the review in the context of what is already known. | 6-7 |
| Objectives | 4 | Provide an explicit statement of questions being addressed with reference to participants, interventions, comparisons, outcomes, and study design (PICOS). | 6-7 |
| METHODS |  |  |  |
| Protocol and registration | 5 | Indicate if a review protocol exists, if and where it can be accessed (e.g., Web address), and, if available, provide registration information including registration number. | N/A |
| Eligibility criteria | 6 | Specify study characteristics (e.g., PICOS, length of follow-up) and report characteristics (e.g., years considered, language, publication status) used as criteria for eligibility, giving rationale. | 4-5 |
| Information sources | 7 | Describe all information sources (e.g., databases with dates of coverage, contact with study authors to identify additional studies) in the search and date last searched. | 8-10 |
| Search | 8 | Present full electronic search strategy for at least one database, including any limits used, such that it could be repeated. | 8 and Appendix 1 |
| Study selection | 9 | State the process for selecting studies (i.e., screening, eligibility, included in systematic review, and, if applicable, included in the meta-analysis). | 9 and Figure 1 |
| Data collection process | 10 | Describe method of data extraction from reports (e.g., piloted forms, independently, in duplicate) and any processes for obtaining and confirming data from investigators. | 10 |
| Data items | 11 | List and define all variables for which data were sought (e.g., PICOS, funding sources) and any assumptions and simplifications made. | 10 |
| Risk of bias in individual studies | 12 | Describe methods used for assessing risk of bias of individual studies (including specification of whether this was done at the study or outcome level), and how this information is to be used in any data synthesis. | 10 |
| Summary measures | 13 | State the principal summary measures (e.g., risk ratio, difference in means). | n/a |
| Synthesis of results | 14 | Describe the methods of handling data and combining results of studies, if done, including measures of consistency (e.g., $I^2$) for each meta-analysis. | 11 |
| Risk of bias across studies | 15 | Specify any assessment of risk of bias that may affect the cumulative evidence (e.g., publication bias, selective reporting within studies). | 10 |
| Additional analyses | 16 | Describe methods of additional analyses (e.g., sensitivity or subgroup analyses, meta-regression), if done, indicating which were pre-specified. | n/a |
| RESULTS |  |  |  |
| Study selection | 17 | Give numbers of studies screened, assessed for eligibility, and included in the review, with reasons for exclusions at each stage, ideally with a flow diagram. | 9-10 |
| Study characteristics | 18 | For each study, present characteristics for which data were extracted (e.g., study size, PICOS, follow-up period) and provide the citations. | 8 and Table 1 and Table 2 |
| Risk of bias within studies | 19 | Present data on risk of bias of each study and, if available, any outcome level assessment (see item 12). | 10-11 |
| Results of individual studies | 20 | For all outcomes considered (benefits or harms), present, for each study: (a) simple summary data for each intervention group (b) effect estimates and confidence intervals, ideally with a forest plot. | 7-8 and table 3 |
| Synthesis of results | 21 | Present results of each meta-analysis done, including confidence intervals and measures of consistency. | 11 |
| Risk of bias across studies | 22 | Present results of any assessment of risk of bias across studies (see Item 15). | 16 |
| Additional analysis | 23 | Give results of additional analyses, if done (e.g., sensitivity or subgroup analyses, meta-regression [see Item 16]). | 14, 17 and Table 4 |
| DISCUSSION |  |  |  |
| Summary of evidence | 24 | Summarize the main findings including the strength of evidence for each main outcome; consider their relevance to key groups (e.g., healthcare providers, users, and policy makers). | 16 |
| Limitations | 25 | Discuss limitations at study and outcome level (e.g., risk of bias), and at review-level (e.g., incomplete retrieval of identified research, reporting bias). | 18-19 |
| Conclusions | 26 | Provide a general interpretation of the results in the context of other evidence, and implications for future research. | 19-20 |
| FUNDING |  |  |  |
| Funding | 27 | Describe sources of funding for the systematic review and other support (e.g., supply of data); role of funders for the systematic review. | n/a |

376 Preferred Reporting Items for Systematic reviews and Meta-Analyses (PRISMA) Checklist

### Supplemental Material #3

RedCap survey: https://redcap.ucsf.edu/surveys/?s=FR93D3Y8K9

### Supplemental Material #3

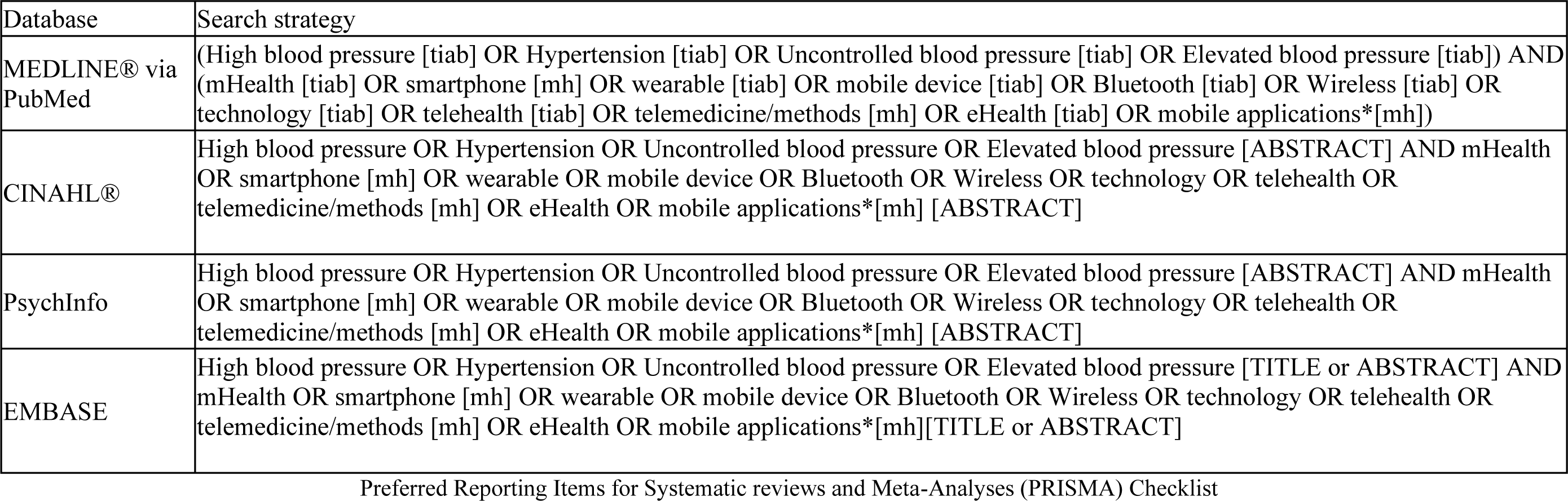

### Supplemental Material #4

**Table 3:**
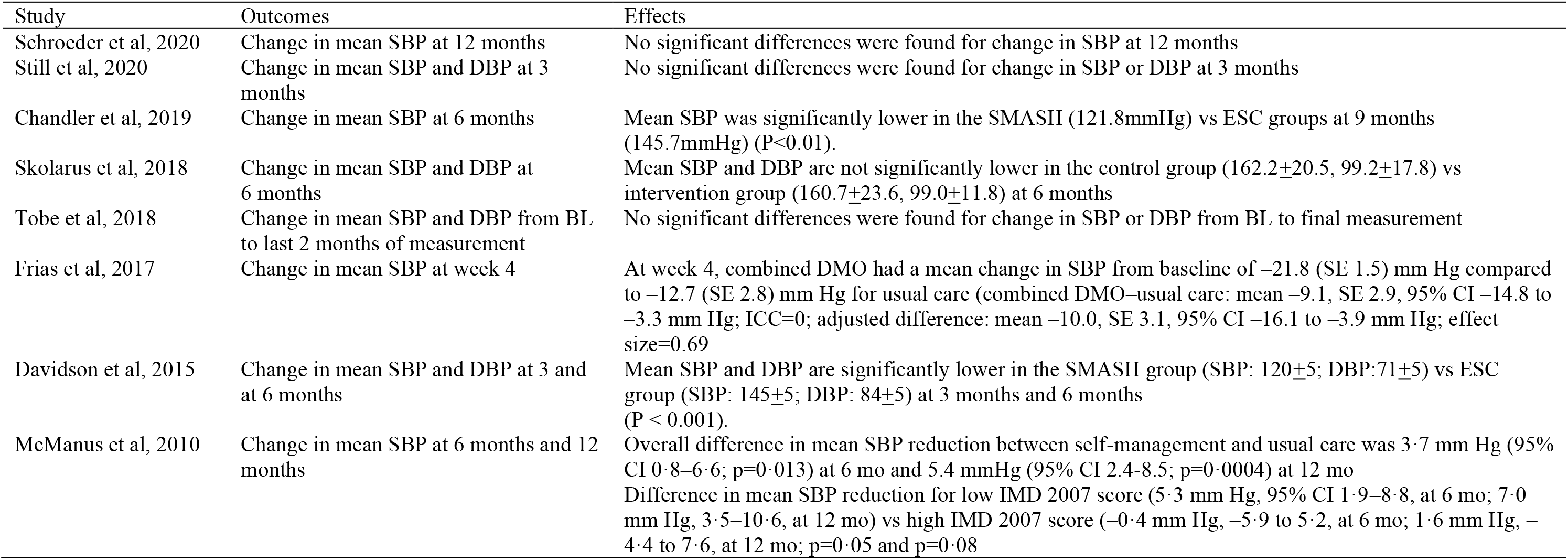
Study Outcomes and Effects.

**Table 4:** Study Quality Assessment via Cochrane Risk of Bias Tool

| Study Information | Random Sequence Generation | Allocation Concealment | Blinding Participants & Personnel | Blinded outcome assessment | Incomplete Outcome Data | Selective Reporting | Other Source of Bias |
| --- | --- | --- | --- | --- | --- | --- | --- |
| Schroeder et al, 2020 | Unknown | Unknown | High | Low | Low | Low | Low |
| Still et al, 2020 | Low | Unknown | High | High | Unknown | Low | Low |
| Chandler et al, 2019 | Unknown | Unknown | Unknown | Unknown | Low | Low | Unknown |
| Skolarus et al, 2018 | Low | Unknown | High | Low | Unknown | Unknown | Unknown |
| Tobe et al, 2018 | Low | Low | High | High | High | High | Unknown |
| Frias et al, 2017 | Low | Low | High | High | Low | Low | Unknown |
| Davidson et al, 2015 | Unknown | Unknown | Unknown | Unknown | Low | Low | Unknown |
| McManus et al, 2010 | Low | High | High | High | Low | Low | Unknown |

